# Hypertension diagnosis, treatment, and control in India: nationally representative results from 1.69 million adults, 2019-2021

**DOI:** 10.1101/2023.06.02.23290909

**Authors:** Jithin Sam Varghese, Nikhil Srinivasapura Venkateshmurthy, Nikkil Sudharsanan, Panniyammakal Jeemon, Shivani A Patel, Harsha Thirumurthy, Ambuj Roy, Nikhil Tandon, KM Venkat Narayan, Dorairaj Prabhakaran, Mohammed K. Ali

**Affiliations:** Emory Global Diabetes Research Center of Woodruff Health Sciences Center and Emory University, Atlanta, USA; Hubert Department of Global Health, Rollins School of Public Health, Emory University, Atlanta, USA; Public Health Foundation of India, New Delhi, India; Professorship of Behavioral Science for Disease Prevention and Health Care, Technical University of Munich, Munich, Germany; Heidelberg Institute of Global Health, Heidelberg University, Germany; Achutha Menon Centre for Health Science Studies, Sree Chitra Tirunal Institute for Medical Sciences and Technology, Trivandrum, India; Leonard Davis Institute of Health Economics and Perelman School of Medicine, University of Pennsylvania, USA; Department of Cardiology, All India Institute of Medical Sciences, New Delhi, India; Department of Endocrinology and Metabolism, All India Institute of Medical Sciences, New Delhi, India; Center for Chronic Disease Control, New Delhi, India; Department of Family and Preventive Medicine, School of Medicine, Emory University, Atlanta, USA

## Abstract

**Background:** Hypertension is a major cause of morbidity and mortality worldwide. Previous efforts to characterize gaps in the hypertension care continuum in India –including diagnosis, treatment, and control– did not assess district level variation. Local data are critical for planning, implementation, and monitoring efforts to curb hypertension burdens. Our objective is to characterize the hypertension care continuum in India among individuals aged 18-98 years old at national, state, and district levels and by socio-demographic group.

**Methods:** Data were from 1,895,297 individuals in the nationally representative Fifth National Family Health Survey (NFHS-5), 2019-21. Hypertension was defined as self-reported diagnosis or newly measured blood pressure ≥140/90 mmHg. Among those with hypertension, we calculated the proportion diagnosed (self-reported). Among those with diagnosed hypertension, we computed the proportion treated (self-reported medication use). Among those treated, we calculated the proportion controlled (BP <140/90 mmHg [20-80 years] or <150/90 mmHg [>80 years]) based on national guidelines. Estimates were also provided among the total with hypertension. To assess differences in the care continuum between or within states (i.e. between districts), we partitioned the variance at both levels using linear mixed models.

**Results:** Among 1,691,109 adult respondents nationally (52.6% female; mean age: 41.6 years), 28.2% [95%CI: 28.0-28.4] had hypertension, of whom, 36.7% [36.3-37.2] were diagnosed. Among those diagnosed, 44.7% [44.1-45.3] reported taking medication (17.7% [17.5-17.9] of total with hypertension). Among those treated, 52.3% [51.4-53.1] had blood pressure control (9.1% [8.9-9.2] of total with hypertension). There were substantial variations across districts in diagnosis [range: 6.3–77.5%], treatment [8.7–97.1%] and control [2.7–76.6%]. Notably, large proportions of the variation in hypertension diagnosis (53.7%), treatment (32.8%), and control (57.7%) were within states, not just between states.

**Conclusions:** In India, more than 1 in 4 people have hypertension, and of these, only 1 in 3 are diagnosed, less than 1 in 5 are treated, and only 1 in 11 controlled. National averages hide considerable state- and district-level variation in the care continuum, implying the need for targeted, decentralized solutions to improve the hypertension care continuum in India.

## Introduction

Hypertension is associated with 12.8% of all deaths globally.^1^ Many countries have implemented large-scale programs to diagnose and manage hypertension and other chronic diseases, with varying success.^2,3^ Of over 1.3 billion people with hypertension globally, 82% live in low- and middle-income countries, and India alone is home to an estimated 220 million adults with hypertension.^4–6^ To address the burden of noncommunicable diseases, India launched the National Programme for Prevention and Control of Cancer, Diabetes, Cardiovascular Disease and Stroke (NPCDCS) in 2010, under the National Health Mission for 100 districts across 21 states.^7^ However, few data are currently available to assess the success and opportunities for improved control of high blood pressure at subnational levels.^8^

Previous efforts to characterize the hypertension care continuum were limited to national and state levels, or exclusively among older or younger adults, but not by socio-demographic groups within states or at district levels.^9–12^ Newer regional data may therefore strengthen ‘planning, implementation, and monitoring of investments’ at the district-level to improve health infrastructure and outreach services for hypertension - key objectives of the Government of India’s national programs.^13–15^

We describe the national, state, and district-level hypertension care continuum (prevalence, diagnosis, treatment, control) in India, the world’s most populous country. We visually represent these data through a publicly available dashboard for stakeholders to help identify priorities for reducing hypertension burdens in India and tracking the progress of national initiatives.

## Methods

### Study Population

The National Family Health Survey-5 (NFHS) is a nationally-representative survey conducted in two phases from June 2019 to March 2020, and from November 2020 to April 2021 in 707 districts from 28 states and 8 union territories, and powered to provide estimates at the district level.^16^ Using a multi-stage stratified approach, primary sampling units (PSUs) were selected from urban (census enumeration blocks) and rural (villages) strata of each district at the first stage. At the second stage, 636,699 households within PSUs were randomly sampled from a list of households where eligible participants (women: 15-49 years, men: 15-54 years) resided.^17^ Household and individual characteristics were collected using standardized instruments. The survey additionally collected data on blood pressure among all adults (18 years and older) who were living in the same household as eligible participants. The overall sample approached consisted of 1,895,297 adults aged 18-98 years.

We restricted our analysis to non-pregnant women and men who had a valid measurement of blood pressure (**Supplementary Figure 1**). The analytic sample consisted of 1,691,109 adults aged 18-98 (47.4% men and 52.6% non-pregnant women), representing a response rate of 89.2%. The analytic sample was similar to the excluded sample (**Supplementary Table 1**). Additional information on sampling and data collected are provided in **Supplementary Methods**.

### Data collection

#### Hypertension

Systolic and diastolic blood pressure were measured three times at five-minute intervals using validated electronic OMRON BP monitors after a five-minute sedentary period when the participant was asked to sit comfortably.^17^ The respondent was also asked to avoid eating, smoking, and exercising for 30 minutes before the measurement. Cuff size of BP monitor was based on circumference of the bare upper arm measured using Gulick tape. Blood pressure was measured on the left arm, positioned so that it was at heart level with the cuff placed over bare skin or over thin clothes. Consistent with 2016 ICMR guidelines, we took the lowest of the first two measurements if their difference in systolic BP was less than or equal to 5 mmHg, and lowest of the three measurements otherwise.^18^

Participants were also asked the question: “Before this survey, were you ever told you had high blood pressure by a doctor, nurse, or health practitioner on two or more occasions?”. Medication status was asked only to those who self-reported a diagnosis of hypertension.

#### Hypertension Care Continuum – Diagnosis, Treatment, and Control

We defined hypertension as self-reported or, among those without a prior diagnosis, measured blood pressure ≥140/90 mmHg.^18^ We defined the hypertension care continuum using the following metrics: proportion diagnosed (self-reported diagnosed hypertension prior to the survey among total with hypertension), and among those diagnosed, the proportion treated (those self-reporting medication use). We defined the proportion controlled among those treated (<140/90 mmHg for those below than 80 years, and <150/90 mmHg for those 80 years and older) based on ICMR guidelines for management of hypertension.^18^ We also provided age-standardized estimates of treatment and control among all of those with hypertension. The definitions are summarized in **Supplementary Table 2**.

#### Socio-demographic variables

We estimated care continuum metrics by 3 individual-level socio-demographic factors: sex (male or female), age (18-39, 40-64, ≥65 years), and schooling (none or missing, primary [up to 4^th^ class], secondary [up to 10^th^ class], post-secondary). We also stratified by two household socio-demographic factors: rural residence (versus urban) and regional wealth quintile (urban and rural) from the household wealth index as provided by NFHS.^19^

### Statistical Analysis

We report survey-weighted estimates accounting for the complex survey design and 95% cluster-robust confidence intervals.^16^ Individual and household characteristics of the analytic sample were assessed by strata of residence (urban or rural) and sex.

Continuum performance indicators were estimated for the national sample, for states stratified by socio-demographic factors (residence, sex, age category, schooling, and regional wealth quintile) and for districts. Age-standardized estimates of the continuum indicators were computed for different strata at the national-level, based on distribution of the total sample since different strata of schooling and wealth have different age distributions. We also calculated weighted estimates at state-level and district-level that were not age-standardized, but would be relevant for local decision making. We compared the estimates to those obtained when taking the average of the last 2 blood pressure measurements as a sensitivity analysis.

To assess whether the differences in the care continuum were greater between or within states (i.e. between districts), we partitioned the variance in the care continuum at both levels using variance partition coefficients from linear mixed models with state-level intercepts. To illustrate the variability between- and within-states, we present examples of two states from regions with moderate to high burdens of hypertension, namely Karnataka from South India and Meghalaya from North East India.

To further aid policy and priority decision-making, we developed a dashboard to visually depict the disparities in the hypertension care continuum using Shiny by RStudio (link provided in the Results). We displayed disparities, both crude and age-standardized, by sex and region (Total/Urban/Rural) at the state-level on the “Overview” tab. We compared districts within each state on the “District Disparities” tab. We displayed disparities across socio-demographic characteristics at the state-level on the “Socio-demographic Disparities” tab. All analyses were carried out using R 4.2.0 using srvyr 1.1.1.

## Results

Nationally, over three-fourths of the population lived in rural areas in 2019-2021. More than half were under 40 years of age and almost 90% were aged 18-64 years (**Table 1**). Average systolic and diastolic BP were 120.3 [95%CI: 120.2-120.4] mmHg and 79.7 [79.7-79.8] mm Hg, respectively, for women, and 124.6 [95%CI: 124.5-124.7] mmHg and 81.7 [81.6-81.7] mm Hg, respectively, for men. (**Table 1**)

**Table 1.** Characteristics of participants in analytic sample for estimating care cascade of hypertension in India, n = 1,691,109

|  | **Total** |  | **Urban** |  | **Rural** |  |
| --- | --- | --- | --- | --- | --- | --- |
|  | **Women**<br>**(n = 889,507)** | **Men**<br>**(n = 801,602)** | **Women**<br>**(n = 218,595)** | **Men**<br>**(n = 198,910)** | **Women**<br>**(n = 670,912)** | **Men**<br>**(n = 602,692)** |
| **Age category** |  |  |  |  |  |  |
| 18-39 | 50.1 (49.9,<br>50.2) | 49.1 (48.9,<br>49.3) | 49.5 (49.1,<br>49.9) | 49.6 (49.3,<br>50.0) | 50.3 (50.2,<br>50.5) | 48.8 (48.6,<br>49.1) |
| 40-64 | 39.4 (39.2,<br>39.5) | 38.7 (38.6,<br>38.9) | 40.3 (40.0,<br>40.5) | 39.2 (38.9,<br>39.5) | 38.9 (38.8,<br>39.1) | 38.5 (38.3,<br>38.7) |
| 65 and above | 10.6 (10.5,<br>10.7) | 12.2 (12.1,<br>12.3) | 10.2 (10.0,<br>10.5) | 11.1 (10.9,<br>11.4) | 10.7 (10.6,<br>10.9) | 12.7 (12.5,<br>12.8) |
| **Schooling** |  |  |  |  |  |  |
| None | 37.1 (36.8,<br>37.4) | 17.4 (17.2,<br>17.6) | 22.3 (21.8,<br>22.8) | 9.5 (9.2, 9.8) | 44.0 (43.7,<br>44.3) | 21.2 (21.0,<br>21.5) |
| Primary (up to 4<sup>th</sup> class) | 13.8 (13.7,<br>14.0) | 15.1 (14.9,<br>15.2) | 12.6 (12.3,<br>12.8) | 11.4 (11.1,<br>11.7) | 14.4 (14.3,<br>14.6) | 16.9 (16.7,<br>17.0) |
| Secondary (5<sup>th</sup> to 10<sup>th</sup> class) | 36.7 (36.5,<br>36.9) | 49.5 (49.3,<br>49.7) | 43.3 (42.9,<br>43.7) | 50.8 (50.3,<br>51.3) | 33.5 (33.3,<br>33.8) | 48.9 (48.6,<br>49.2) |
| Post-secondary (11<sup>th</sup> class and above) | 12.4 (12.2,<br>12.6) | 18.0 (17.7,<br>18.3) | 21.8 (21.3,<br>22.3) | 28.3 (27.7,<br>28.9) | 8.0 (7.9, 8.2) | 13.0 (12.7,<br>13.4) |
| **Household wealth quintile**<br>**(by residence)** |  |  |  |  |  |  |
| Lowest | 18.6 (18.3,<br>18.9) | 18.0 (17.6,<br>18.3) | 19.4 (18.6,<br>20.1) | 19.4 (18.6,<br>20.1) | 18.2 (17.9,<br>18.6) | 17.3 (17.0,<br>17.6) |
| Low | 19.6 (19.4,<br>19.8) | 19.3 (19.1,<br>19.6) | 20.2 (19.7,<br>20.7) | 20.2 (19.7,<br>20.7) | 19.3 (19.1,<br>19.5) | 18.9 (18.6,<br>19.2) |
| Medium | 20.3 (20.1,<br>20.5) | 20.4 (20.2,<br>20.6) | 20.3 (19.9,<br>20.8) | 20.3 (19.9,<br>20.8) | 20.3 (20.1,<br>20.5) | 20.4 (20.2,<br>20.7) |
| High | 20.7 (20.4,<br>20.9) | 21.2 (20.9,<br>21.4) | 20.3 (19.8,<br>20.8) | 20.3 (19.8,<br>20.8) | 20.9 (20.6,<br>21.1) | 21.6 (21.3,<br>21.9) |
| Highest | 20.8 (20.5,<br>21.2) | 21.1 (20.8,<br>21.5) | 19.9 (19.2,<br>20.6) | 19.7 (19.0,<br>20.5) | 21.3 (20.9,<br>21.7) | 21.8 (21.5,<br>22.2) |
| **Blood pressure measurement** |  |  |  |  |  |  |
| Average Systolic BP (mmHg) | 120.3 (120.2,<br>120.4) | 124.6 (124.5,<br>124.7) | 120.6 (120.4,<br>120.8) | 125.4 (125.2,<br>125.6) | 120.2 (120.1,<br>120.3) | 124.2 (124.1,<br>124.3) |
| Average Diastolic BP (mmHg) | 79.7 (79.7, 79.8) | 81.7 (81.6, 81.7) | 79.9 (79.8, 80.0) | 82.1 (82.0, 82.3) | 79.6 (79.5, 79.7) | 81.5 (81.4, 81.5) |
| **Hypertension** |  |  |  |  |  |  |
| Self-reported or high blood pressure | 27.2 (27.0, 27.4) | 28.4 (28.2, 28.7) | 29.1 (28.7, 29.5) | 31.1 (30.6, 31.5) | 26.3 (26.1, 26.5) | 27.2 (26.9, 27.4) |
| Self-reported | 12.6 (12.5, 12.8) | 9.1 (9.0, 9.3) | 15.0 (14.6, 15.3) | 11.2 (10.9, 11.5) | 11.6 (11.4, 11.8) | 8.1 (8.0, 8.3) |
| **Blood Pressure Category** |  |  |  |  |  |  |
| BP <120/80 mm Hg | 40.1 (39.9, 40.3) | 27.3 (27.1, 27.6) | 38.3 (37.9, 38.7) | 24.7 (24.3, 25.1) | 41.0 (40.8, 41.2) | 28.5 (28.3, 28.8) |
| 120/80 ≤ BP < 140/90 mmHg | 39.7 (39.5, 39.9) | 48.7 (48.5, 48.9) | 40.6 (40.2, 41.0) | 49.5 (49.0, 49.9) | 39.2 (39.0, 39.4) | 48.3 (48.1, 48.5) |
| 140/90 ≤ BP < 160/100 mm Hg | 14.4 (14.3, 14.5) | 17.7 (17.6, 17.9) | 15.4 (15.1, 15.7) | 19.2 (18.8, 19.5) | 14.0 (13.8, 14.1) | 17.0 (16.8, 17.2) |
| BP ≥ 160/110 mmHg | 5.8 (5.7, 5.9) | 6.3 (6.2, 6.4) | 5.7 (5.6, 5.9) | 6.6 (6.4, 6.8) | 5.8 (5.7, 5.9) | 6.1 (6.0, 6.2) |

All values are percentages (95% confidence intervals) accounting for survey design. Estimates are not age-standardized. The household wealth index, as provided by the Demographic and Health Surveys, was computed as the first principal component from survey responses regarding possession of assets and quality of housing, separately for urban and rural areas.

### National-level care continuum

The age-standardized prevalence [95%CI] of hypertension (**Table 2**) nationally was 28.2% [95%CI: 28.0, 28.4] and was higher in urban areas (32.7% [32.3-33.1]) relative to rural areas (25.9% [25.7-26.2]). The prevalence was higher among men (30.6% [30.4, 30.9]) relative to women (25.8% [25.6-26.0]), and was higher at older ages (65 and above: 54.3% [53.8-54.8], 18-39: 15.0% [14.9-15.2]), and greater household wealth (highest: 31.2% [30.8-31.5], lowest: 25.5% [25.2-25.8]) compared to their respective counterparts. Higher hypertension prevalence in men, older, and wealthier individuals was observed in both urban and rural areas. Prevalence of hypertension did not vary by education at the national level.

**Table 2.** Socio-demographic variations in care continuum in India, n = 1,691,109

|  | **Total** |  |  |  | **Urban** |  |  |  | **Rural** |  |  |  |
| --- | --- | --- | --- | --- | --- | --- | --- | --- | --- | --- | --- | --- |
|  | **Hyperten**<br>**sion (%)** | **Diagnose**<br>**d<sup>a</sup> (%)** | **Treated<sup>b</sup>**<br>**(%)** | **Controlle**<br>**d<sup>c</sup> (%)** | **Hyperten**<br>**sion (%)** | **Diagnose**<br>**d<sup>a</sup> (%)** | **Treated<sup>b</sup>**<br>**(%)** | **Controlle**<br>**d<sup>c</sup> (%)** | **Hyperten**<br>**sion (%)** | **Diagnose**<br>**d<sup>a</sup> (%)** | **Treated<sup>b</sup>**<br>**(%)** | **Controlle**<br>**d<sup>c</sup> (%)** |
| **Total** | 28.2<br>(28, 28.4) | 36.7<br>(36.3, 37.2) | 44.7<br>(44.1, 45.3) | 52.3<br>(51.4, 53.1) | 32.7<br>(32.3, 33.1) | 39.8 (39,<br>40.7) | 56.3<br>(54.9, 57.6) | 50.2<br>(48.7, 51.7) | 25.9<br>(25.7, 26.2) | 35.2<br>(34.7, 35.8) | 38.8 (38,<br>39.6) | 53.7<br>(52.6, 54.7) |
| **Sex** |  |  |  |  |  |  |  |  |  |  |  |  |
| Women | 25.8<br>(25.6, 26) | 44.4<br>(43.8, 44.9) | 42.2<br>(41.5, 42.9) | 55.3<br>(54.3, 56.4) | 30.1<br>(29.7, 30.6) | 47.7<br>(46.7, 48.8) | 54 (52.5,<br>55.5) | 52.9<br>(51.1, 54.6) | 23.8<br>(23.6, 24) | 42.9<br>(42.2, 43.6) | 36.9 (36,<br>37.8) | 56.9<br>(55.6, 58.2) |
| Men | 30.6<br>(30.4, 30.9) | 28.3<br>(27.9, 28.8) | 49.3<br>(48.5, 50.1) | 47.2<br>(45.8, 48.5) | 35.2<br>(34.7, 35.7) | 32.1<br>(31.3, 33) | 59.9<br>(58.3, 61.5) | 46.3 (44,<br>48.6) | 28.3 (28,<br>28.5) | 26.2<br>(25.7, 26.8) | 42.8<br>(41.8, 43.9) | 47.8<br>(46.2, 49.5) |
| **Age**<br>**category** |  |  |  |  |  |  |  |  |  |  |  |  |
| 18-39 | 15 (14.9,<br>15.2) | 31.3<br>(30.6, 32) | 23.8<br>(22.9, 24.7) | 60.8<br>(59.2, 62.4) | 15.7<br>(15.3, 16) | 28.4<br>(27.1, 29.7) | 27.2<br>(25.3, 29) | 57.1<br>(54.1, 60) | 14.8<br>(14.6, 15) | 32.4<br>(31.6, 33.2) | 22.7<br>(21.7, 23.7) | 62.9<br>(61.1, 64.7) |
| 40-64 | 37.2 (37,<br>37.5) | 39.5 (39,<br>39.9) | 61.8<br>(61.1, 62.4) | 43.7 (43,<br>44.3) | 40.2<br>(39.7, 40.7) | 44.4<br>(43.6, 45.2) | 70 (68.8,<br>71.2) | 44.6<br>(43.5, 45.6) | 35.5<br>(35.2, 35.7) | 36.5 (36,<br>37) | 56 (55.2,<br>56.8) | 42.9<br>(42.2, 43.7) |
| 65 and<br>above | 54.3<br>(53.8, 54.8) | 51.3<br>(50.7, 51.9) | 77.1<br>(76.5, 77.8) | 44.3<br>(43.5, 45.1) | 60.1<br>(59.1, 61.1) | 59.8<br>(58.6, 60.9) | 83.9<br>(82.8, 85) | 45.8<br>(44.4, 47.2) | 50.5 (50,<br>51) | 45.6 (45,<br>46.3) | 71.7<br>(70.8, 72.5) | 43.2<br>(42.3, 44.1) |
| **Schooling** |  |  |  |  |  |  |  |  |  |  |  |  |
| None | 27.7<br>(27.5, 28) | 36.2<br>(35.5, 36.9) | 41.6<br>(40.7, 42.6) | 47.4<br>(46.2, 48.5) | 32.9<br>(32.2, 33.6) | 38.8<br>(37.2, 40.5) | 52.1<br>(49.4, 54.7) | 44.5 (42,<br>46.9) | 26.6<br>(26.3, 26.9) | 35.5<br>(34.7, 36.3) | 38.8<br>(37.7, 40) | 48.4 (47,<br>49.8) |
| Primary<br>(up to 4<sup>th</sup><br>class) | 28.8<br>(28.5, 29.1) | 35.9<br>(35.2, 36.6) | 46.8<br>(45.6, 48) | 50.7 (49,<br>52.4) | 34.8 (34,<br>35.5) | 38.2<br>(36.8, 39.6) | 58.9<br>(56.5, 61.3) | 46.3<br>(43.6, 49) | 26.8<br>(26.4, 27.1) | 35 (34.1,<br>35.8) | 42 (40.6,<br>43.5) | 53 (50.9,<br>55.2) |
| Secondary<br>(5<sup>th</sup> to 10<sup>th</sup><br>class) | 28.2 (28,<br>28.4) | 36.9<br>(36.4,<br>37.4) | 45.8<br>(45.1,<br>46.5) | 52.8<br>(51.4,<br>54.1) | 33 (32.5,<br>33.5) | 39.9 (39,<br>40.8) | 57.9<br>(56.4,<br>59.4) | 49.6<br>(47.6,<br>51.7) | 25.4<br>(25.2,<br>25.7) | 35 (34.4,<br>35.6) | 37.9<br>(36.9,<br>38.8) | 55.1<br>(53.4,<br>56.9) |
| Post-secondary<br>(11<sup>th</sup> class<br>and above) | 28.3<br>(27.8,<br>28.7) | 39.3<br>(38.4,<br>40.2) | 46.9<br>(45.7,<br>48.1) | 59.4<br>(56.9, 62) | 31.1<br>(30.4,<br>31.8) | 42 (40.7,<br>43.2) | 55.4<br>(53.5,<br>57.3) | 57 (53.5,<br>60.4) | 24.5<br>(23.9,<br>25.1) | 35.6<br>(34.5,<br>36.7) | 34.6 (33,<br>36.3) | 63.3<br>(59.6,<br>66.9) |
| **Household**<br>**wealth**<br>**quintile** |  |  |  |  |  |  |  |  |  |  |  |  |
| Lowest | 25.5<br>(25.2,<br>25.8) | 31.9 (31,<br>32.7) | 37.2 (36,<br>38.4) | 54.5<br>(52.5,<br>56.5) | 29.1<br>(28.4,<br>29.8) | 34.8 (33,<br>36.5) | 45.1<br>(42.5,<br>47.7) | 49.9<br>(46.7, 53) | 23.6<br>(23.3, 24) | 30.2<br>(29.2,<br>31.2) | 32.2<br>(30.6,<br>33.7) | 58.2<br>(55.5,<br>60.9) |
| Low | 26.8<br>(26.5,<br>27.1) | 35.7 (35,<br>36.4) | 41.9<br>(40.9,<br>42.9) | 51.4<br>(49.6,<br>53.2) | 32 (31.4,<br>32.7) | 38.1<br>(36.8,<br>39.4) | 55.3<br>(53.2,<br>57.5) | 47.3<br>(44.5, 50) | 24 (23.7,<br>24.3) | 34.4<br>(33.5,<br>35.3) | 34 (32.7,<br>35.3) | 55 (52.8,<br>57.3) |
| Medium | 27.9<br>(27.6,<br>28.2) | 36.7 (36,<br>37.4) | 44.5<br>(43.4,<br>45.5) | 52.1<br>(50.3,<br>53.9) | 33.3<br>(32.7, 34) | 39.6<br>(38.4,<br>40.8) | 59.6<br>(57.6,<br>61.7) | 50.3<br>(47.4,<br>53.2) | 25.2<br>(24.9,<br>25.5) | 35.2<br>(34.3,<br>36.1) | 36.5<br>(35.2,<br>37.8) | 53.5<br>(51.2,<br>55.9) |
| High | 29.1<br>(28.8,<br>29.4) | 37.1<br>(36.4,<br>37.9) | 47.2<br>(46.2,<br>48.3) | 52.7<br>(50.9,<br>54.4) | 34 (33.3,<br>34.6) | 41.6<br>(40.3,<br>42.9) | 59.3<br>(57.2,<br>61.4) | 51 (48,<br>54) | 26.7<br>(26.4,<br>27.1) | 35.1<br>(34.2, 36) | 41.4 (40,<br>42.7) | 53.7<br>(51.7,<br>55.8) |
| Highest | 31.2<br>(30.8,<br>31.5) | 40.6<br>(39.8,<br>41.3) | 48.1 (47,<br>49.2) | 51.7 (50,<br>53.4) | 34.8<br>(34.1,<br>35.5) | 44.2<br>(42.7,<br>45.7) | 58.7<br>(56.3, 61) | 51.6<br>(48.3,<br>54.9) | 29.5<br>(29.1,<br>29.9) | 39.1<br>(38.2,<br>39.9) | 43.8<br>(42.4,<br>45.1) | 51.7<br>(49.9,<br>53.6) |

Estimates (95% confidence intervals) are standardized to age distribution in overall sample.
a Among those with self-reported hypertension or high blood pressure ( $\geq 140/90$ mmHg).
b Among those with self-reported hypertension ('Diagnosed')
c Among those taking medication for hypertension ('Treated')

Among all adults with hypertension, 36.7% [36.3-37.2] reported being diagnosed, (**Figure 1**). Diagnosed hypertension was higher in urban (39.8% [39.0-40.7]) compared to rural areas (35.2% [34.7-35.8]), higher among older age groups (65 and above: 51.3% [50.7-51.9], 18-39: 31.3% [30.6-32.0]), and those with greater household wealth (highest: 40.6% [39.8-41.3], lowest: 31.9% [31.0-32.7]) but did not vary by schooling (post-secondary: 39.3% [38.4-40.2], none: 41.6% [40.7-42.6]).

**Figure 1.**
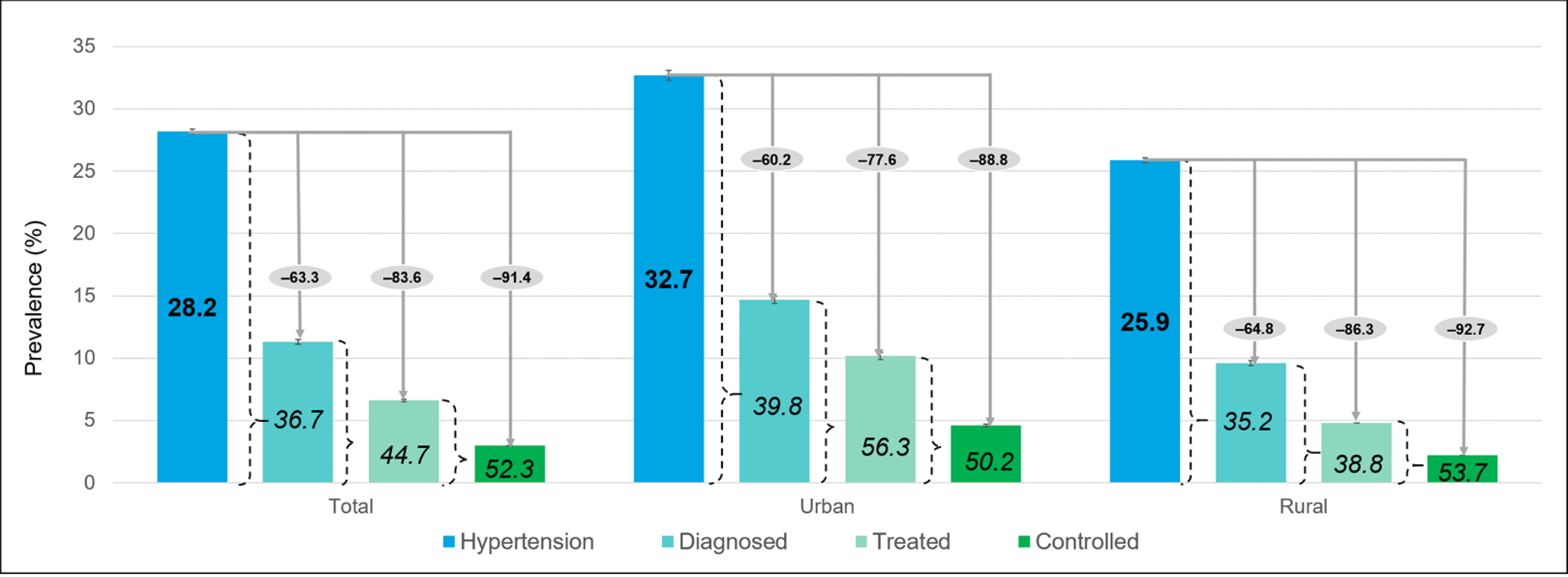
National-level care continuum in Indian adults by residence, n = 1,691,109. All columns are survey weighted percentages in total population. The values in *italics* are proportions of diagnosed hypertension among patients with hypertension, treated among diagnosed hypertension and controlled among treated hypertension (from **Table 2**). We performed age-standardization to the distribution of the within-sample total population separately for total population, population with hypertension, diagnosed population and treated population. This procedure harmonizes the age distribution within each category (total, hypertension, diagnosed, treated). The values should therefore not be sequentially multiplied to get prevalence within total population. Values in grey ovals are drops from all patients with hypertension (100 - %diagnosed among hypertension, 100 - %treated among hypertension, 100 - %controlled among hypertension).

Of adults with diagnosed hypertension, 44.7% [44.1-45.3] reported taking medication corresponding to 17.7% [17.5-17.9] of those with hypertension (**Figure 1; Supplementary Table 3**). Among those diagnosed, medication use was 56.3% [54.9-57.6] in urban areas and 38.8% [38.0-39.6] in rural areas. These estimates correspond to 23.9% [23.4, 24.4] and 14.6% [14.4, 14.9] of those with hypertension. Proportions of those diagnosed that were treated was higher among men (men: 49.3% [48.5-50.1], women: 42.2% [41.5-42.9]), with higher age (65 and above: 77.1% [76.5-77.8], 18-39: 23.8% [22.9-24.7]) and household wealth (lowest: 37.2% [36.0-38.4], highest: 48.1% [47.0-49.2]), but did not vary by education. The distributions of those treated and controlled, treated and uncontrolled, or untreated, among those diagnosed, are presented in **Supplementary Figure 2**. Estimates of treated and controlled hypertension among those with hypertension by socio-demographic group are provided in **Supplementary Table 3**.

Among those diagnosed and treated with medication, 52.3% [51.4-53.1] had controlled blood pressure corresponding to 9.1% [8.9-9.2] of all those with hypertension (**Figure 1; Supplementary Table 3**). Among treated adults, the proportion with controlled hypertension was 50.2% [48.7-51.7] in urban areas and 53.7% [52.6-54.7] in rural areas. These estimates correspond to 7.6% [7.4-7.8] and 12.0% [11.7-12.4] of those with hypertension. Controlled hypertension among those treated was higher among women (55.3% [54.3-56.4]) than men (47.2% [45.8-48.5]), and adults aged 18-39 years (60.8% [59.2-62.4]) compared to 40-64 years (43.7% [43.0-44.3]) and those older than 65 years (44.3% [43.5-45.1]). Hypertension control was also higher with higher schooling (none: 47.4% [46.2-48.5], post-secondary: 59.4% [56.9-62.0]), but did not differ by household wealth (lowest: 54.5% [52.5-56.5], medium: 52.1% [50.3-53.9], highest: 51.7% [50.0-53.4]).

Our results were similar when using the average of the second and third measurements of blood pressure (**Supplementary Table 4**), instead of the lowest measurements (**Supplementary Table 5, Supplementary Figure 3**).

### State-level care continuum

Hypertension prevalence was similar among the southern states (Kerala, Tamil Nadu, Karnataka, Telangana, Andhra Pradesh), union territories (Andaman & Nicobar Islands, Lakshadweep, Puducherry), and Goa compared to other parts of the country (**Figure 2**; median of states: 30% [southern] vs 26.9% [rest of India]). Higher hypertension prevalence was observed in urban versus rural areas for all states (**Supplementary Figure 4**).

**Figure 2.**
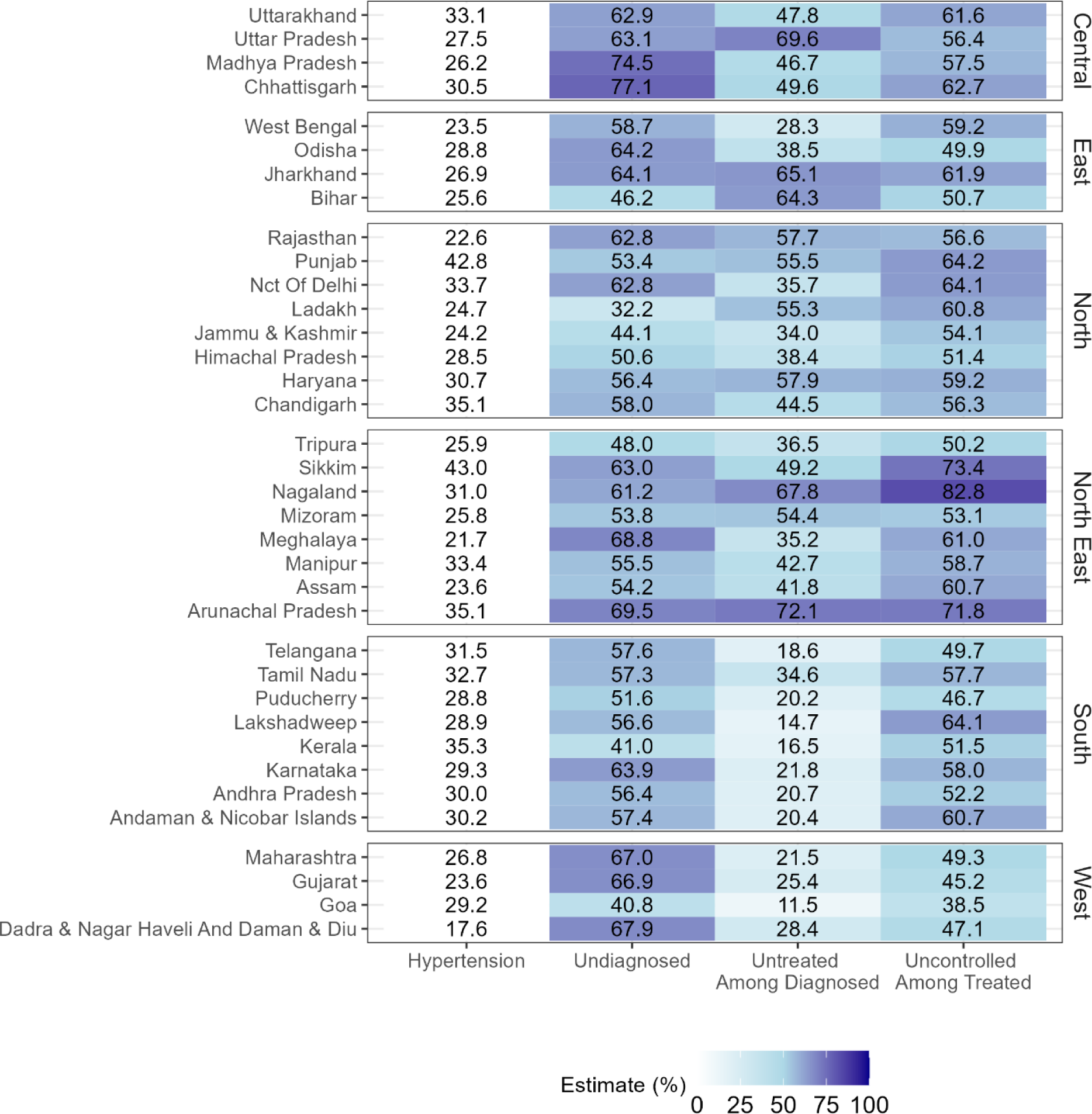
State-level unmet need in hypertension care continuum, n = 1,691,109. All values are survey weighted percentages (not age standardized). Undiagnosed are among those with hypertension. Untreated and uncontrolled are among those diagnosed with hypertension and among those treated respectively. We report weighted estimates at state-level that were not age-standardized and relevant for local decision making in this manuscript. We present age standardized comparisons in the interactive dashboard.

The proportions with diagnosed hypertension were similar between southern states and rest of India (**Supplementary Figure 5)**. However, proportions treated and controlled were higher among the southern states. Disparities in diagnosis, treatment, and control between socio-demographic groups within each state beyond the state-level heterogeneity observed in **Figure 2** are published on the interactive ‘Hypertension Care Continuum’ dashboard (accessed at: https://egdrc-precision-medicine.shinyapps.io/hypertension_cascade/).

### District-level care continuum

There was considerable within-state (between-district) variation in the hypertension care continuum **(Figure 3)** such that 53.7% of variance in proportion diagnosed, 32.8% of variance in proportion treated among diagnosed, and 57.7% of variance in proportions controlled among treated were at the district-level, with the remaining at the state-level (between-state). We visualized this variability between- and within-states from all regions in **Supplementary Figure 6**.

**Figure 3.**
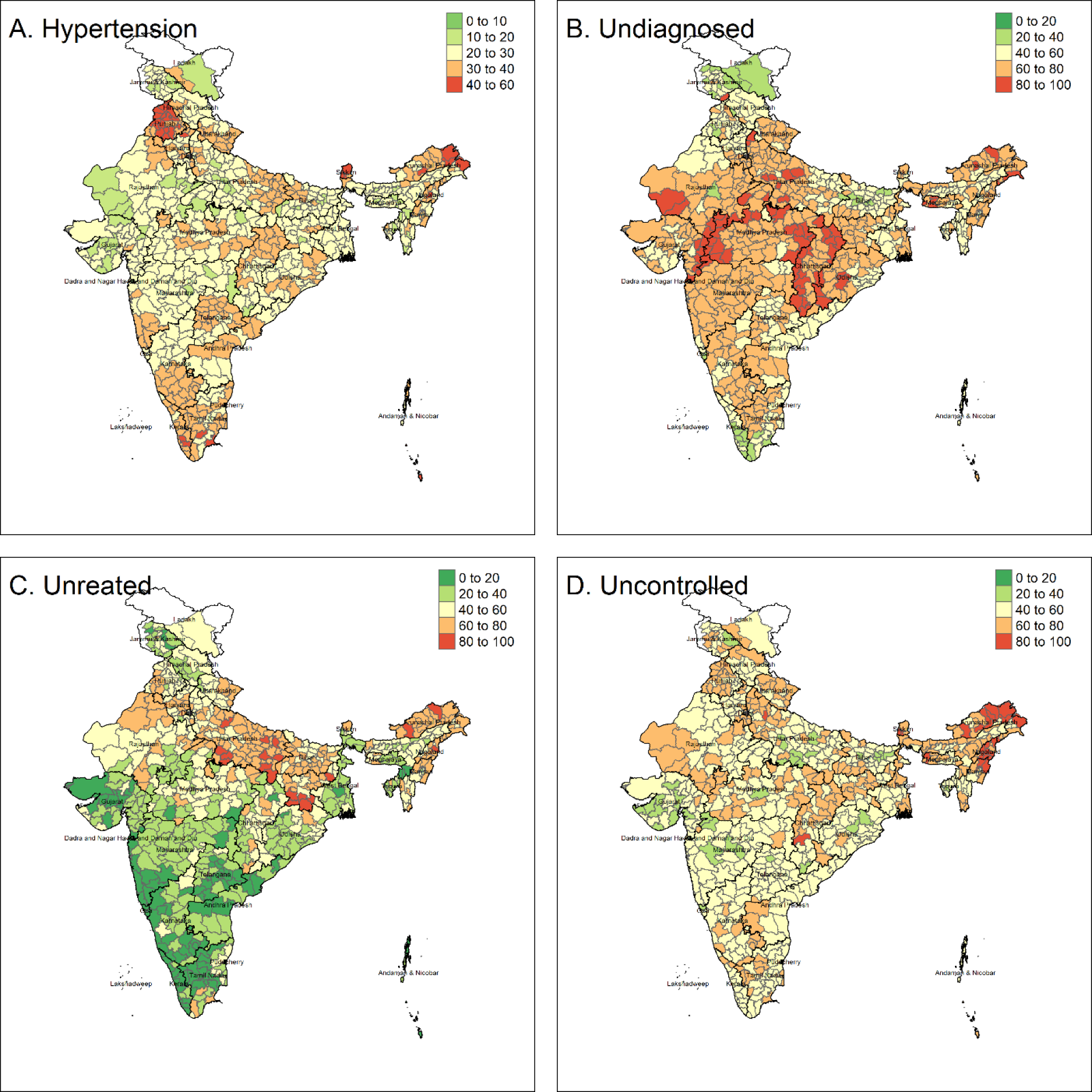
Care continuum in analytic sample by urban and rural residence for 707 districts, n = 1,691,109. All values are survey weighted percentages (not age standardized). Undiagnosed are among those with hypertension. Untreated and uncontrolled are among those diagnosed with hypertension and among those treated respectively. We report weighted estimates at the district-level that were not age-standardized and relevant for local decision making in this manuscript. We present age standardized comparisons in the interactive dashboard.

We illustrated this variability in Meghalaya and Karnataka. In Meghalaya, the five districts of Garo Hills (median: 21.9%) had similar prevalence as the two districts of Jaintia Hills (median: 18.8%) and three districts of Khasi Hills (median: 23.2%) although the proportions of those diagnosed, were much lower in Garo Hills (18.5%) than Jaintia Hills (40.8%) and Khasi Hills (29.4%)(**Supplementary Figure 7A)**.

In Karnataka, there was substantial between-district heterogeneity in treatment among those diagnosed but less heterogeneity in control between districts with similar prevalence. Chikmagalur (31.6% [28.8-34.4]), Udupi (34.0% [31.5-36.6]), Chitradurga (34.8% [32.1-37.5]), and Shimoga (34.0% [29.0-39.0]) had similar prevalence of hypertension. The proportions treated were higher in Chikmagalur (81.3% [84.5-88.0]) and Udupi (91.3% [87.6-95.1]), compared to Chitradurga (61.6% [45.9-77.2]) and Shimoga (55.7% [38.1-73.3]) (**Supplementary Figure 7B)**. Similarly, the proportion of those with controlled hypertension (**Supplementary Figure 7C**) was higher in Chikmagalur (42.5% [33.6-51.4]) and Udupi (44.1% [39.8-48.4]) compared to Chitradurga (39.6% [30.8-48.4]) and Shimoga (35.0% [28.6-41.5]).

## Discussion

Of the estimated 28% adults older than 18 years with high blood pressure in India, nearly 2 in 3 remain undiagnosed across all states and in both urban and rural areas.^20^ Among those diagnosed, only half were treated; treatment was higher in southern and western India, and lower in other parts of the country. Among those treated, nearly half did not have their blood pressure under control. Cumulatively, over 90% of adults with hypertension in India were either undiagnosed, untreated, or treated and uncontrolled.

There was substantial variability across socio-demographic groups in prevalence, diagnosis, treatment, and control of hypertension. Although the prevalence of hypertension was higher among men, the proportion of those diagnosed was higher among women.^11,21^ Women were less likely to be taking medication, but again, treated women were more likely than treated men to have controlled hypertension.^10,11^ Proportion diagnosed did not vary with schooling,^11^ but proportion of treated and controlled were higher among those with higher schooling. Proportion diagnosed and treated were higher among older adults and wealthier households.^10,11^

The reasons for greater differences in hypertension diagnosis, treatment, and control being between districts in a state, and not between states are likely multifactorial. Prior and recent data show that there are between-district differences in health-seeking behaviors across India.^16^ Furthermore, clinician (e.g. type of provider and practice variation) and system (e.g. physical and financial access to clinics) factors also differ between states and districts.^22,23^

The high unmet need in hypertension diagnoses in India has been identified previously, though none of these provide comprehensive estimates for all age groups and district-level precision in estimates.^12,24^ In 2017-18, the National Non-communicable Diseases Monitoring Survey (NNMS) surveyed 10,659 adults aged 18-69y (n=10,659) from 26 states and estimated a hypertension prevalence of 28.5%.^11^ Among those with hypertension, 27.9% were diagnosed, 14.5% were treated (52.0% among diagnosed), and 12.6% were controlled (86.9% among those treated). Although NNMS provided estimates for socio-demographic groups at the national level, they did not provide state-level estimates. The NFHS-4 and Longitudinal Aging Study in India (LASI), both of which were conducted over 2017-19, provided estimates for those aged 15-49 years (n=731,864; prevalence: 18.1%, diagnosed: 44.7%, treated among diagnosed: 29.8%, controlled among treated: 59.4%) and those older than 45 years (n=72,262; prevalence: 45.9%, diagnosed: 55.7%, treated among diagnosed: 69.8%, controlled among diagnosed: 56.9%), respectively.^9,10,25^ NFHS-4 and LASI provided estimates by socio-demographic group and state.^14,15^

To improve the care continuum for hypertension in India, our data suggest that diagnosis is a critical step in realizing the downstream indicators such as treatment and control. Screening and linkage to care are therefore critical, as evidenced by previous data.^26–28^ Studies within India also offer promising opportunities to improve hypertension diagnosis by linking frontline health workers who carry out hypertension screening at the community level with doctors at the facility level though an IT-enabled platform.^29^ In addition, under the Ayushman Bharat Comprehensive Primary Healthcare (CPHC) program for screening and referral for non-communicable diseases, digitization of screening records by frontline workers can enable surveillance of hypertension burdens.^30^ Concerted strategies for hypertension treatment and control may offer models for India to emulate.^31–33^ Hypertension control can also be facilitated by providing doctors latest evidence-based guidelines on treating hypertension through decision support systems embedded within the NPCDCS portal.^29^ Furthermore, population-based strategies such as policy mandated reductions in salt content of packaged foods, food labeling, low sodium or salt substitutes,^34^ reducing particulate exposure, and improved built environments can complement the above-mentioned clinical efforts.

Our study has key strengths. This study is among the largest of its kind, consisting of over 1.6 million respondents, providing data at district level, and for sociodemographic groups. The response rate was high, and there were few missing data, indicating high quality of data collection and completion. The study used validated protocols for blood pressure measurement including cuff size selection, and our presentation offers easy-to-use visualization of results.

Our study had some limitations. First, while the hypertension care continuum is an invaluable tool to visualize gaps at one time-point, these data hide the dynamic nature of hypertension treatment and control, and argues for systems of ongoing surveillance.^35,36^ Second, hypertension, among those who did not self-report a physician diagnosis, was based on blood pressure measurements at a single time point.^37^ The ICMR guidelines for diagnosis of hypertension requires a minimum of 2 sets of readings on 2 different occasions, which are at least 1-4 weeks apart.^18^ Third, diagnosis and treatment were based on self-report, and not validated through medical records.^35,38^ Finally, we did not have data on older adults living by themselves or institutionalized and non-civilian adults.^16^

In India, nationally, more than 1 in 4 people have hypertension, and cumulatively, over 90% of adults with hypertension were either undiagnosed, untreated, or treated and uncontrolled. These summary data, however, hide district-level and sociodemographic differences. Thus, as our data indicate, the characterization and visualization of India’s hypertension care continuum nationally, at the state and district levels, and across socio-demographic groups present opportunities to tailor implementation of programs to prevent and control the burdens of high blood pressure.

## Data Availability

All datasets used in this analysis are available for download at www.dhsprogram.com. The code for the analysis is available on https://github.com/jvargh7/hypertension_cascade.

https://dhsprogram.com/

## Abbreviations

ICMR: Indian Council of Medical Research
LASI: Longitudinal Aging Study in India
NFHS: National Family Health Surveys
NNMS: National Noncommunicable Disease Monitoring Survey
UT: Union Territory
WHO: World Health Organization

## Acknowledgements

We thank the participants and survey enumerators of National Family Health Survey 2019-21.

